# Unraveling the Role of Viral Interference in Disrupting Biennial RSV Epidemics in Northern Stockholm

**DOI:** 10.1101/2024.08.09.24310749

**Authors:** Ke Li, Johan Hamrin, Daniel M. Weinberger, Virginia E. Pitzer

## Abstract

Respiratory syncytial virus (RSV) primarily impacts infants and older adults, with seasonal winter outbreaks in temperate countries. Biennial cycles of RSV activity have also been identified in Northern Europe and some states in the United States. Delayed RSV activity was reported worldwide during the 2009 influenza pandemic, and a disrupted biennial pattern of RSV activity was observed in northern Stockholm following the pandemic. Biennial patterns shifted to early/large outbreaks in even-numbered years and late/small outbreaks in odd-numbered years. However, the mechanisms underpinning this change in pattern remain unknown. In this work, we constructed an age-stratified mechanistic model to explicitly test three factors that could lead to the change in RSV transmission dynamics: 1) birth rates, 2) temperatures, and 3) viral interference. By fitting the model to weekly RSV admission data over a 20-year period and comparing different models, we found that viral interference from influenza was the only mechanism that explained the shifted biennial pattern. Our work demonstrates the complex interplay between different respiratory viruses, providing evidence that supports the presence of interactions between the H1N1 pandemic influenza virus and RSV at the population level, with implications for future public health interventions.

## Introduction

Respiratory syncytial virus (RSV) infections are a major public health concern for pediatric populations, older adults, and immunocompromised individuals [1,2]. In 2019, an estimated 100,000 deaths of children under the age of 5 were attributed to RSV globally [3]. RSV is highly seasonal, with winter epidemics in temperate countries. Biennial cycles of RSV activity have also been identified in Northern Europe and some states in the United States.[4–9] In northern Stockholm, RSV activity shows a regular biennial pattern with early and large epidemics in odd-numbered seasons (e.g., 2001/02 season), and late and small epidemics in even-numbered seasons (e.g., 2002/03 season), influencing the risk of both lower respiratory infection and hospitalizations [10,11].

Disruptions to RSV epidemics were observed worldwide following the 2009 influenza pandemic, and delayed RSV activity was reported in the 2009/10 season [12–17]. In north Stockholm, our observations revealed an unexpected annual pattern for RSV epidemics during 2009-2011, characterized by large epidemics in two consecutive seasons. In particular, the biennial pattern of RSV epidemics completely shifted to early and large seasons on even-numbered years following the pandemic season. This disruption to the previous patterns provides an opportunity to understand factors that influence the annual and biennial epidemic cycles of RSV. To explain the change of RSV activity in northern Stockholm, we propose three hypotheses: 1) a sudden increase of birth rates in 2009, 2) extremely low temperatures during the winters of 2009/10 and 2010/11, and 3) the occurrence of the 2009 influenza pandemic, which may interfere with RSV activity.

Mathematical models that explicitly depict the underlying mechanisms of virus transmission have advantages in being able to integrate heterogeneous mechanisms and test different hypotheses [18,19]. In this work, we started by analyzing weekly admission data for RSV from 1998 to 2018 in north Stockholm and demonstrated when and how RSV activity was disrupted in the area. We then constructed a mechanistic, age-stratified mathematical model that allows us to investigate different hypotheses for why the RSV biennial pattern may have shifted. Applying both a maximum likelihood method and a sampling-importance-resampling method, we estimated climatic and viral interference parameters and compared different models based on the hypotheses. Finally, we used the best-fit model to predict RSV dynamics under different scenarios, explaining how the number of susceptible individuals impacted RSV transmission.

## Results

### Disruption of temporal patterns of RSV

Hospitalizations for RSV, from July 1998 to June 2018, were strongly seasonal and showed an annual pattern of outbreaks occurring during the winter months in northern Stockholm (**Fig. 1A and 1B**). A biennial pattern was also detected, with early/large RSV epidemics in odd-numbered years (e.g., 1999/00 and 2001/02 years, highlighted in shaded areas, **Fig. 1A**) and late/small epidemics in even-numbered years (e.g., 2002/03 and 2000/01 years, **Fig. 1A**) prior to 2009. In odd-numbered years prior to the 2009 influenza pandemic, the mean timing (as indicated by the center of gravity) and the mean value of RSV activity intensity (as indicated by the peak value of RSV hospitalizations) was 31.2 weeks and 30.6 cases, respectively, compared to 35.7 weeks and 19.2 cases in even-numbered years (**Fig. S1**). However, we observed that the temporal pattern was disrupted during the 2009/10 and 2010/11 seasons. The timing of the biennial cycle exhibited a consistent pattern before 2009, but there was a sudden change in timing from 2009 to 2011 (**Fig. 1C**). The pattern shifted to early/large epidemics in even-numbered years and to late/small epidemics in odd-numbered years (highlighted in the shaded area, **Fig. 1A**) from 2010 to 2018. The timing of the annual cycle was consistent over time. These patterns can be quantified using the phase angle, which exhibits a shift in timing for the biennial cycles around 2009 Following the pandemic, the mean timing and the mean value of RSV activity intensity are 35.2 weeks and 22.8 cases in odd-numbered years, respectively, compared to 31.5 weeks and 32 cases in even-numbered years (**Fig. S1**).

**Figure 1.**
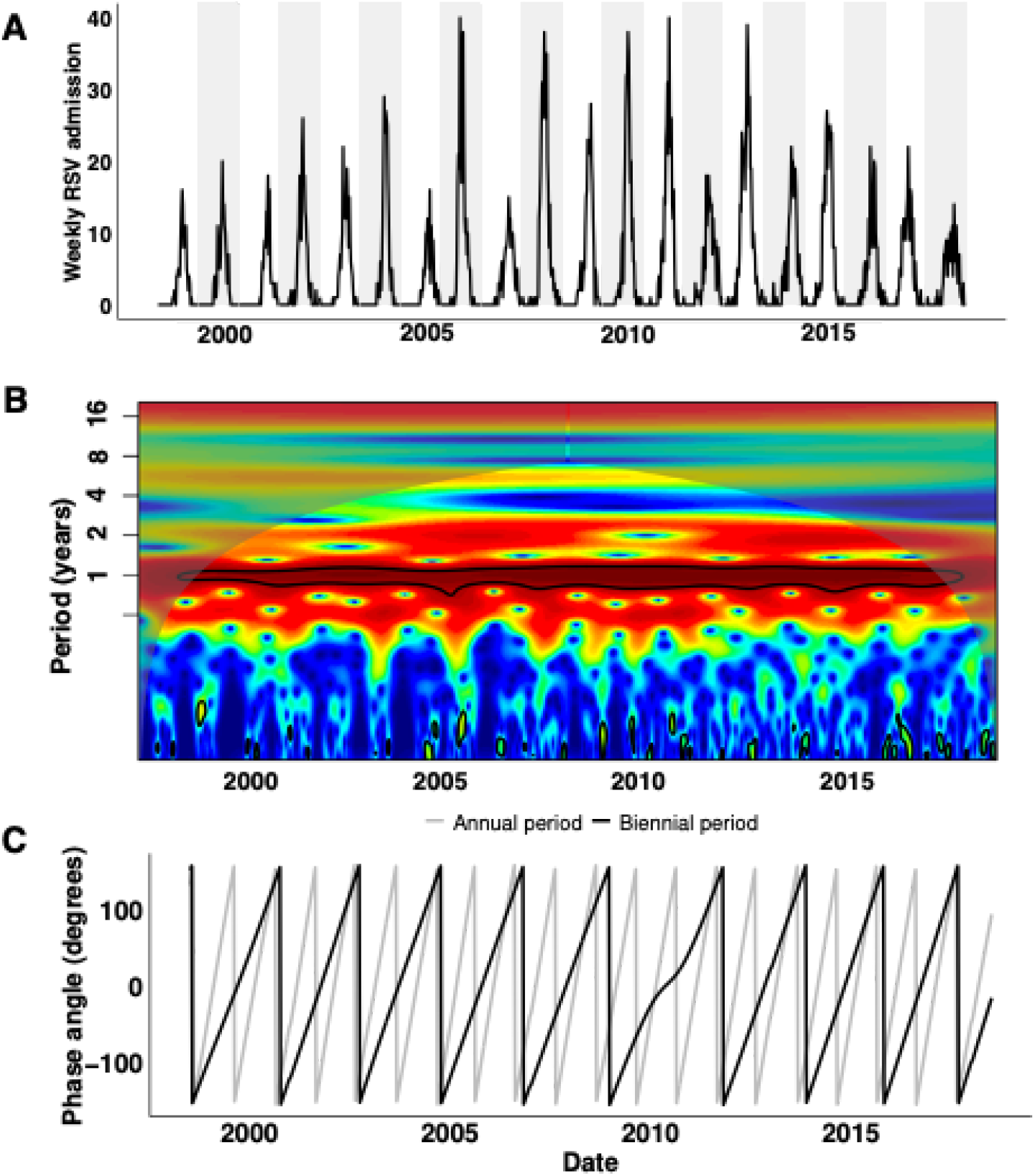
RSV epidemics in northern Stockholm. (**A**) Weekly hospital admissions due to RSV from July 1998 to June 2018 from the catchment area of Astrid Lindgren Children’s Hospital in northern Stockholm. Odd-numbered seasons are highlighted in shaded areas. (**B**) The period (in years) of RSV epidemics. (**C**) The phase angle (in degrees) of the annual period (gray) and biennial period (black) of RSV epidemics.

### Hypotheses for disrupted RSV dynamics

Having detected and quantified the disruption of RSV activity, we then sought to explore what factors led to the pattern change. The disrupted period (i.e., 2009-2011 years) coincided with three notable changes. The first observation was a sudden increase in the birth rate. By analyzing the annual birth rate data in northern Stockholm, we found that there was a 10% increase in the birth rate from 2010 to 2011 (**Fig. S2A**). Second, we observed that the area experienced exceptionally cold weather during the winters of 2010 and 2011 (**Fig. S2B**). Third, the occurrence of the H1N1 influenza pandemic during the 2009-2010 season (**Fig. S2C**). Therefore, we hypothesized that the three changes may be associated with the disruption of RSV activity.

### Dynamic model analyses

To explore the mechanistic relationship between these factors and the changes in the biennial pattern, we built an age-stratified SIS (Susceptible-Infectious-Susceptible) model for RSV transmission dynamics based upon the model from Pitzer et al. [20], accounting for repeat infections (**Fig. 2**) and using natural history parameters derived from RSV cohort studies (**S1 Table**). See **Materials and Methods** for a detailed model description.

**Figure 2.**
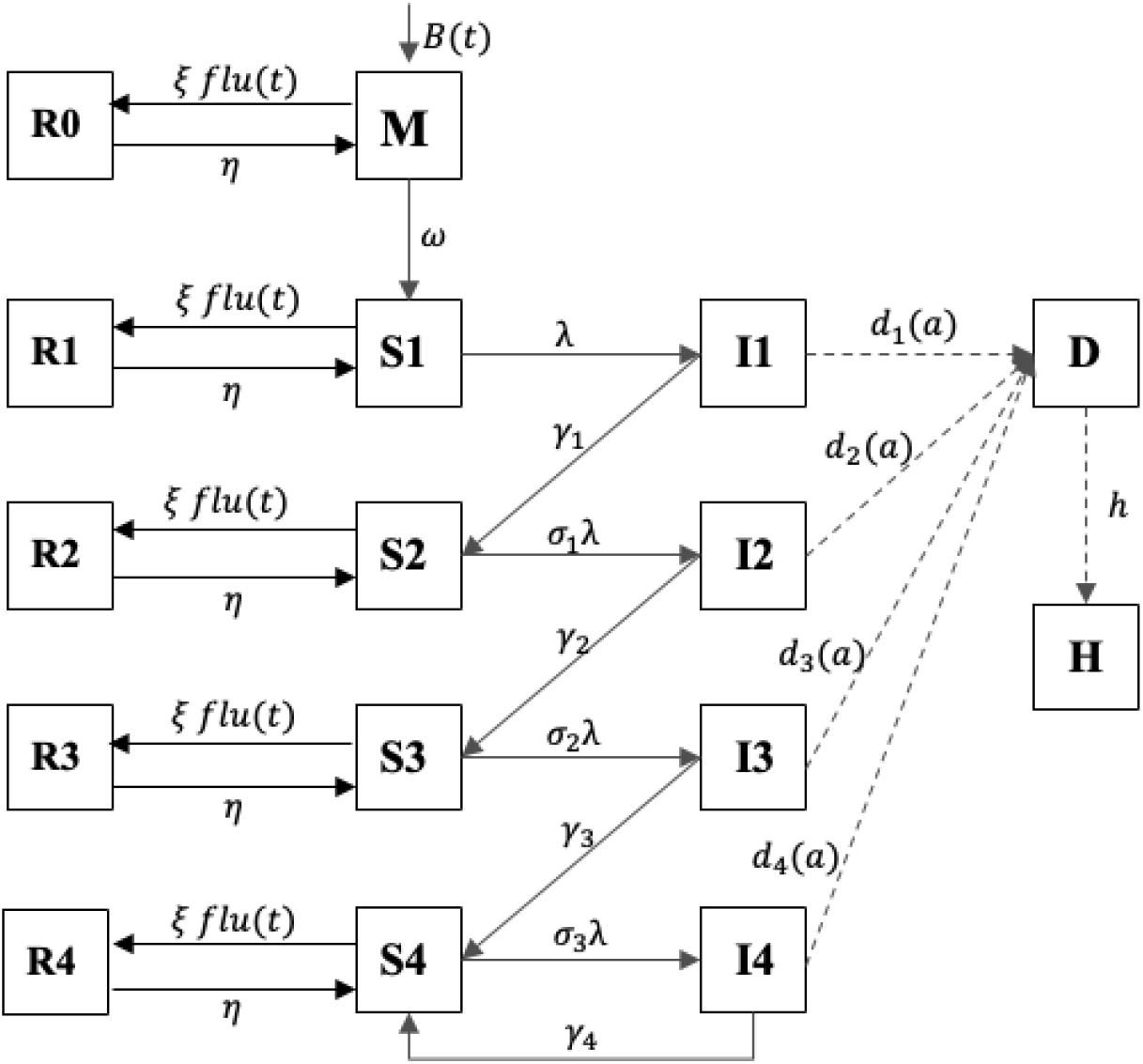
Transmission dynamic model for RSV. Model diagram illustrating the structure of the RSV transmission model (age is omitted for simplicity). Components *S*1*-S*4represent susceptible populations; components *I*1*-I*4represent infected populations, and components *R*0*-R*4represent temporarily-immune populations. The population with maternal antibodies is represented by *M*. Components *D* and *H* represent the population that develops severe lower respiratory infections and that is hospitalized, respectively. Note that the two components are not infection states in the model, and they represent observed states. See **S1 Text** for detailed model equations.

To explore whether the sudden increase in birth rates explains the biennial pattern change, we first calibrated the RSV transmission model to weekly RSV admissions from 1998 to 2008 (i.e., the seasons prior to the 2009 influenza pandemic) using a maximum likelihood method to estimate key parameters governing RSV transmission (see **Materials and Methods**). We then forward-simulated the model with the estimated parameters. We found that although the model could reproduce the biennial pattern before the disputed period (i.e., 1999-2009), it could not capture the altered biennial pattern of RSV epidemics after the pandemic season (i.e., 2009-2019, **Fig. 3A**). Using the same approach, we incorporated the time-series of normalized temperatures and relative humidity data into the model, and fitted it to weekly RSV admissions. Again, we found that the model with climatic factors could not capture the disputed RSV epidemics following the influenza pandemic (**Fig. 3B**).

**Figure 3.**
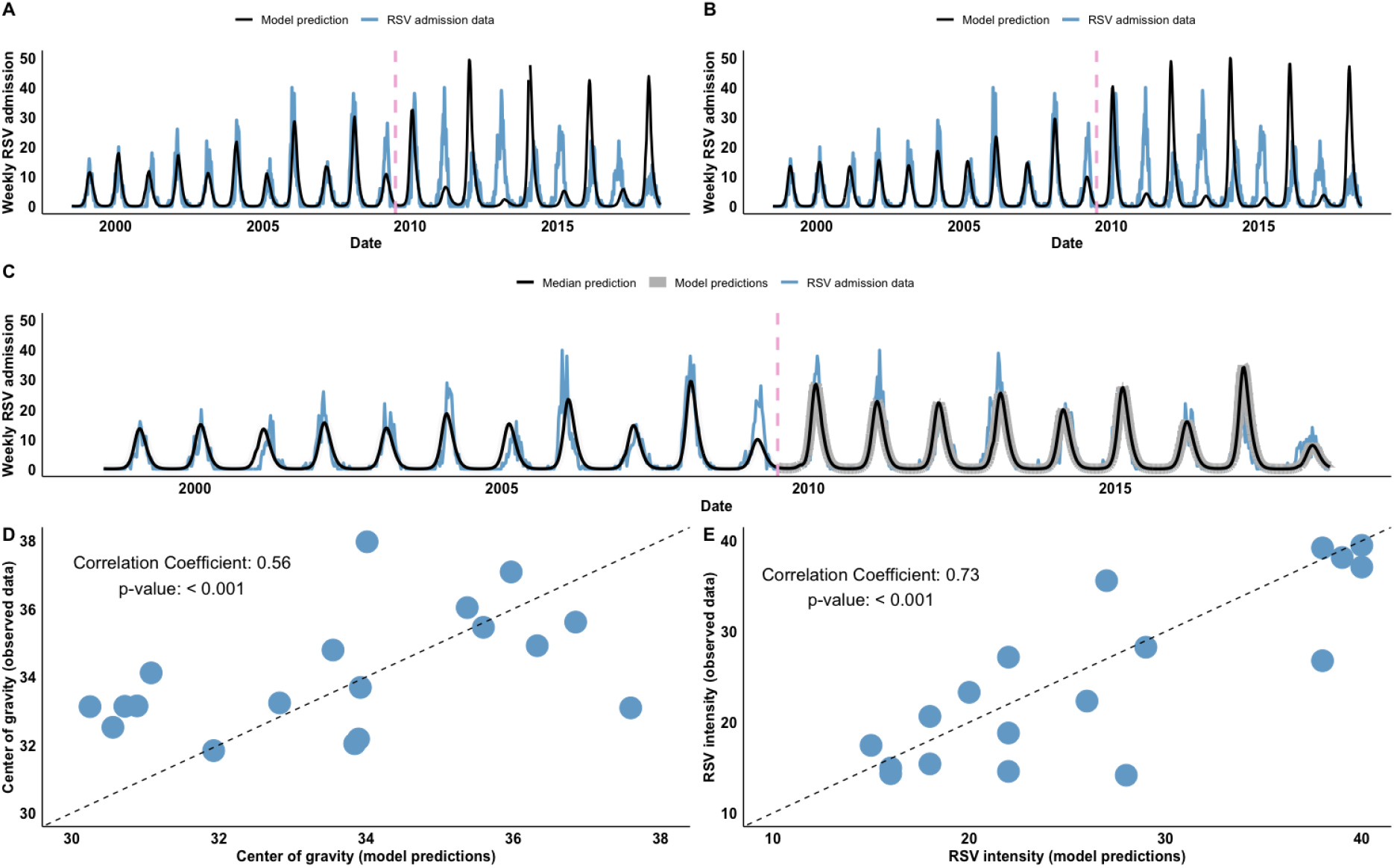
Model fit to age-stratified RSV admissions to test different hypotheses. (**A**) Model fit to weekly RSV data, assuming no viral interference effects (i.e., ξ=0, see **S2 Table** for estimated parameter values). (**B**) Model fit to weekly RSV, temperature and relative humidity data, assuming no viral interference effects (i.e., ξ=0, see **S3 Table** for estimated parameter values). (**C**) The number of observed weekly RSV admissions are shown in blue; the median model prediction, given by the median estimates of viral interference parameters, is shown in black, and other randomly selected model predictions are shown in gray. The pandemic H1N1 influenza virus was introduced to the model only in the 2009/10 season, and the pink dashed line indicates July 2009. Correlations between (**D**) the center of gravity (in weeks) and (**E**) the intensity of observed and predicted seasonal RSV epidemics from the best-fit model with the viral interference effect.

Next, we sought to investigate whether viral interference from influenza explains the change in the biennial pattern of RSV transmission. For this, we introduced temporary immune populations into the model (i.e., *R*0*-R*4, **Fig. 2**). We assumed that the temporary protection was mounted by influenza infection through the host innate immune response [21]. Here, we did not explicitly model the transmission dynamics for pandemic influenza due to increased model complexity. Instead, we used weekly influenza admissions data as model inputs, and assumed that the conversion rate from susceptible (i.e., *S*1*-S*4) to be temporarily immune was proportional to the number of influenza admissions.

To estimate the effect of viral interference (i.e., ξ) and the duration of temporary protection (i.e., η), we implemented a sampling-importance resampling method (see **Materials and Methods** for details). With viral interference effects from influenza, we found that the model could successfully capture the observed RSV dynamics (**Fig. 3C**). The model was able to reproduce the shifted biennial pattern following the 2009/10 pandemic season, as observed in the data, showing late/small epidemics in odd-numbered years and early/large epidemics in even-numbered years. Notably, the model predicted a larger epidemic for the 2010/11 season as observed. We further calculated the intensity and mean timing of RSV activity of each epidemic season for both observed and model-predicted data from different models. For the models without the viral interference effect, we found negative correlations between the observed and predicted center of gravity (**Fig. S3A, S3C**) and between the observed and predicted RSV intensity (**Fig. S3B, S3D**), indicating poor alignment between the data and model predictions. Instead, we found a positive correlation between the observed and predicted center of gravity of RSV epidemics (**Fig. 3D**, Pearson’s correlation coefficient *r* = 0.56, *p* < 0.001), and between the observed and predicted RSV intensity (**Fig. 3E**, *r* = 0.73, *p* < 0.001). The results showed that the model with viral interference effects well represented the observed RSV dynamics and the shifted biennial patterns, suggesting viral interference from influenza had a significant role in impacting RSV transmission. We further investigated if incorporating climate factors into the viral interference model improved the model fit. However, we found that including temperature and relative humidity data did not enhance the model fit (**Fig. S4**).

### Parameter estimates suggest viral interference

The identified marginal distributions for viral interference parameters provided insight into the effect of viral interference (i.e., ξ) and the duration of temporary protection (i.e., η) from influenza against RSV infection (**Fig. 4**). The median estimate for the viral interference effect parameter (on a log10-scale) was -1.80, with a 95% credible interval (CI) [-2.20, -1.12] (**Fig. 4A**). The median estimate for the duration of temporary immunity was 6.78 days with a 95% CI [1.46, 16.10] (**Fig. 4B**). The results suggest the existence of a viral interference effect from pandemic influenza that contributed to the disrupted biennial pattern of RSV epidemics. Notably, we observed a nonlinear negative correlation (*r =* 0.95, *p <* 0.001) between the two viral interference parameters (**Fig. 4C**). We also found that the sampled parameter sets were more concentrated around and reached the highest density at an effect size of 0.03 per influenza case per week, and at the duration of protection of 3.76 days. The results indicated a short-lived protection provided by host immunity against subsequent RSV infection.

**Figure 4.**
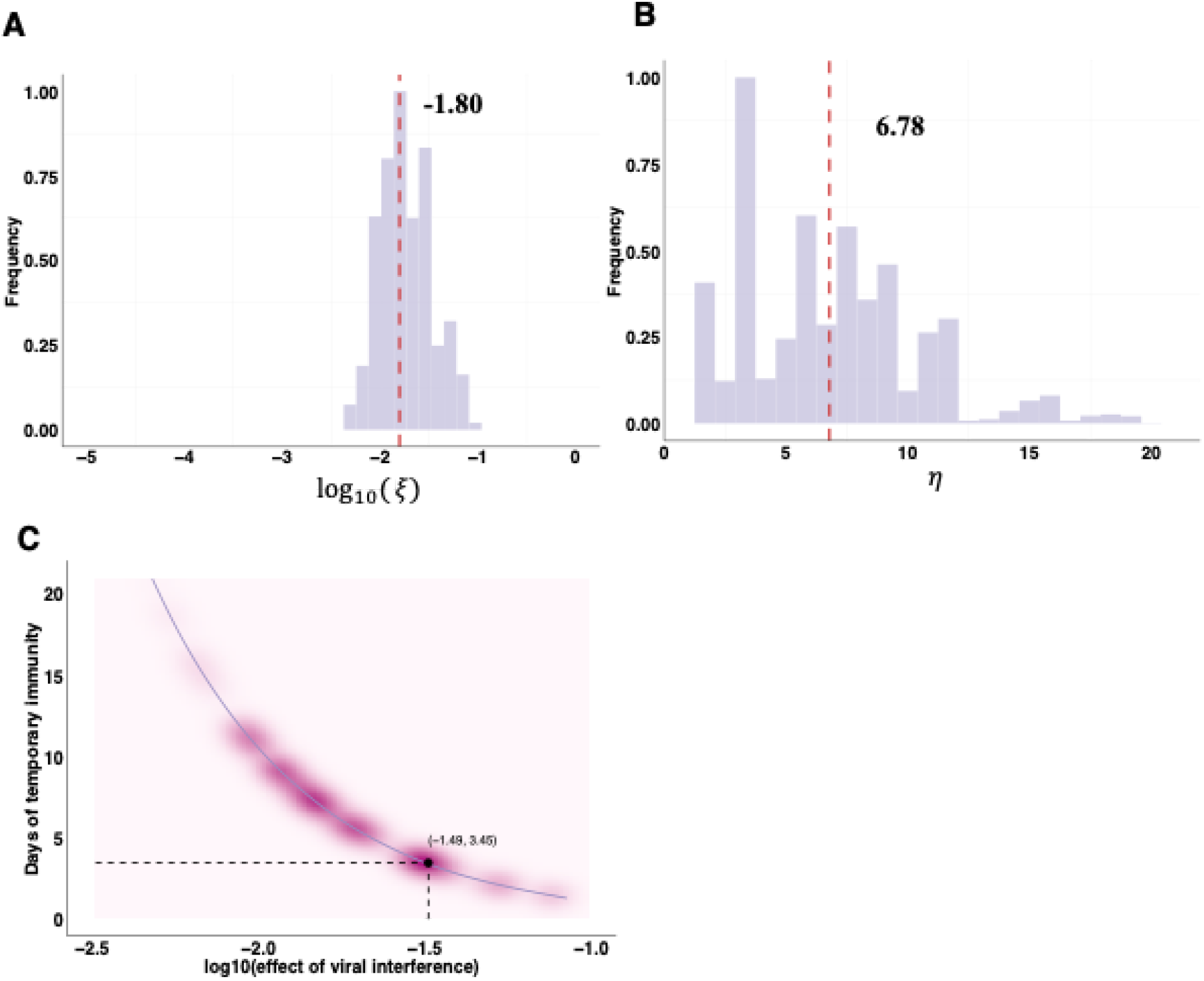
Estimated parameter space for viral interference parameters. (**A**) The marginal distribution for the effect of viral interference parameter (ξ, on a log10-scale). The median estimate is indicated by a red dashed line. (**B**) The marginal distribution for the duration of viral interference parameter (η). (**C**) Correlation between the estimated effects and the duration of viral interference. The parameter set with the highest density is represented by the black point.

### Linking susceptible population dynamics to RSV epidemics

Having demonstrated that the change of the RSV biennial pattern can be explained by viral interference from the pandemic H1N1 influenza, we then sought to understand how viral interference disrupted RSV epidemics. For this, we aggregated model-predicted time series of susceptible populations from 2008 to 2012 (i.e., *S1*, **Fig. 5**; *S2-S4* see **Figs. S5-S7**) of all age groups from the best-fitting model with and without the viral interference effect, respectively. We found a temporal change of susceptible populations in the presence of viral interference. The temporary conversion of the susceptible population (i.e., *S1*, **Fig. 5A**) to the temporarily-immune population (i.e., *R1*, **Fig. 5A**) delayed the availability of susceptible individuals for RSV infection, consequently delaying an RSV epidemic in the 2009/10 season (**Fig. 5B**). This also provided an explanation for a large epidemic in the 2010/11 season, as there were more susceptible individuals after the pandemic season. Furthermore, we observed that the altered susceptible population dynamic did not revert to its pre-pandemic pattern, leading to the shifted biennial pattern for RSV transmission (**Fig. 5B**).

**Figure 5.**
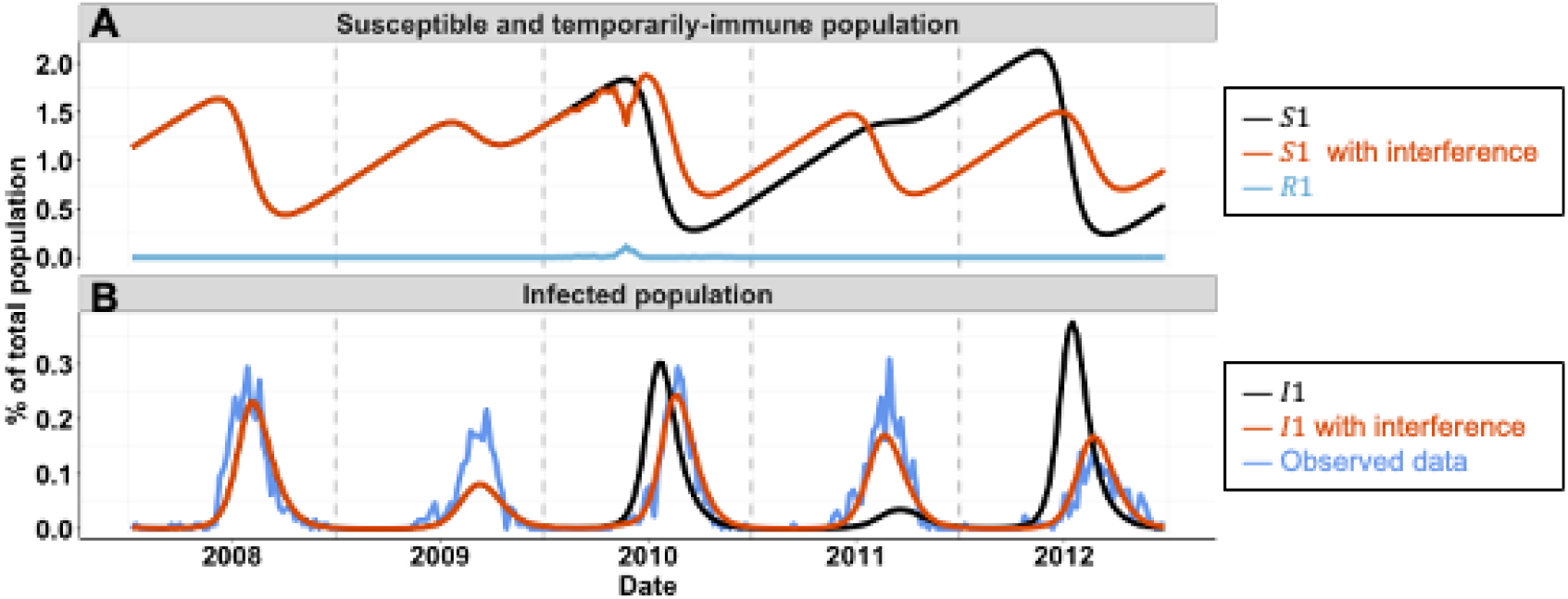
Model predictions of RSV epidemics following the influenza pandemic. (**A**) Predicted time series of susceptible populations in the absence (black curve) or in the presence (red curve) of viral interference effects. The predicted time series of the temporarily immune population is shown in blue. (**B**) Predicted time series of infected populations in the absence (black curve) or in the presence (red curve) of viral interference effects. Observed data are shown in blue. Predicted time series of recovered susceptible populations (i.e., *S2-S4*) and reinfected susceptible populations (i.e., *I*2*-I*4) are provided in **Figs. S5-S7**.

## Discussion

By analyzing the unexpected disruption of the biennial pattern of RSV epidemics in northern Stockholm, we found evidence supporting the presence of immunological interactions between the 2009 H1N1 pandemic influenza virus and RSV [21,22] at the population level. Using an age-stratified dynamic transmission model for RSV, we assessed three hypotheses for the shift in the biennial pattern of RSV. We demonstrated that the sudden rise in birth rates and unusually cold temperatures in 2009/10 and 2010/11 could not explain the disruptions in RSV activity. Instead, only the model incorporating viral interference effects could successfully reproduce the observed RSV pattern change following the 2009 influenza pandemic, implying a significant role of viral interference from influenza in influencing RSV transmission. By applying both a maximum likelihood method and a sampling-importance resampling method, we estimated viral interference parameters, showing the presence of viral interference from influenza on subsequent RSV infection for a short period of time. In our previous work, we also demonstrated the presence of viral interference effects from influenza that impacted RSV epidemics following the 2009 influenza pandemic in the United States [23]. We further analyzed the model-predicted time series of susceptible populations, revealing that the change in the biennial pattern of RSV could be explained by temporary disruptions in the susceptible population.

Mechanistic models provide a valuable approach for dissecting underlying causal relationships among different components, integrating heterogeneous mechanisms and studying various hypotheses. The presence of viral interaction between different viruses is evident at the host level [24–28]. In particular, within-host effects of viral interference from the pandemic H1N1 virus to subsequent RSV infection has been studied using a ferret model [21]. Here, by utilizing a mechanistic model and fitting it to the dataset capturing unusual disruptions of the biennial pattern of RSV, we demonstrated the impact of viral interference on RSV transmission at the population level. One key finding from the model estimates is that temporary protection from viral interference to subsequent RSV infection is transient, lasting less than a week. This finding is consistent with an *in vivo* study [21], indicating that viral interference effects were only observed when the time interval between primary and challenge infections was less than 7 days.

Changes in birth rates have been shown to affect infectious disease dynamics. Models that incorporate changes in birth rates and vaccination levels can explain the complex transition from annual epidemics to irregular or multi-year cycles in measles incidence [29]. More generally, Morris et al. have shown that for diseases with high rates of loss of immunity, a change in birth rate will have negligible impact on the timing of epidemics [30]. Longer-term variations in birth rates may help explain changing patterns of RSV epidemics, such as the transition from a biennial pattern to an annual pattern during the 2000s in California, United States [20]. By contrast, our results showed that a temporary increase of birth rates in 2009 was not sufficient to explain the shift in the biennial pattern of RSV activity in northern Stockholm. Although there may be a delayed and/or long-term impact of birth rates on RSV dynamics, more work is needed to understand the causal relationship between birth rates and RSV transmission.

Using statistical or mathematical models, several studies have examined the relationship between climatic factors and RSV seasonality, and a significant association was found between temperature [31–36], potential evapotranspiration [20], vapor pressure [20], precipitation [20], and relative/absolute humidity [34,37] and RSV activity in different geographic areas. In particular, Baker et al. demonstrated that local climate changes would influence the future dynamics of RSV epidemics, and the timing and intensity of RSV outbreaks would vary by location, influenced by the actual changes in climate [38]. Here, our results showed that the fluctuation of climatic variables, including temperatures and relative humidity, were not key factors responsible for the back-to-back large RSV epidemics in 2009/10 and 2010/11 seasons in northern Stockholm. Climatic factors alone could not reproduce the observed RSV pattern, and incorporating them into the viral interference model did not improve its ability to explain the data. It is possible that extreme temperatures in a single season might temporarily affect the timing of RSV activity, but they are unlikely to have a lasting impact on RSV transmission. Additional studies across different geographic settings could provide more insights into how different climatic factors affect RSV activity.

There are some limitations to our study. First, we did not explicitly model RSV-A and RSV-B in this work due to data scarcity in northern Stockholm. It is possible that the interaction between RSV types A and B may help to explain some of the RSV transmission dynamics, as shown in [39]. However, differences in the predominant circulating subtype of RSV is unlikely to be the main driver of biennial patterns of RSV transmission [5,40,41]. Second, we could not differentiate between genetic subtypes of influenza viruses due to data limitations, and our model only took into account influenza cases in the 2009/10 season, assuming all admissions were due to infection with the pandemic H1N1 influenza virus. This assumption is reasonable, because the pandemic H1N1 influenza virus circulated as the dominating influenza virus in the 2009/10 season in Sweden (96.9% of 51,000 tested samples were pandemic H1N1), and only influenza B was detected in the 2010/11 season [42]. We assumed that viral interference occurs only with the pandemic H1N1 virus and not with other A subtypes of influenza or influenza B. This assumption is based on the fact that the innate immunity is stimulated differently by the pandemic influenza virus compared to other seasonal influenza viruses (see review [40]). Third, we did not have age information on the weekly admissions for RSV and influenza. Therefore, we assumed a well-mixed population and did not account for varying levels of immunity across different age groups beyond the age-specific contact matrices. We also tested a different model structure in which we assumed that only older age groups could be temporarily protected by influenza infection, but the model did not fit the data better.

Understanding the impact of the dynamics of susceptible populations on the transmission of infectious diseases is critical for predicting and preparing for future outbreaks. By studying the unique biennial pattern change in northern Stockholm, our study demonstrated the complex interplay between different respiratory viruses and the implications for public health interventions. We highlighted that even a temporary, small disruption of the susceptible population caused by cross-protection mediated by viral interference effects led to a change in RSV activity. The delay in the RSV epidemic during the 2009/10 season was attributed to a temporary reduction in susceptible individuals, while the subsequent increase in susceptibility led to a larger epidemic in the 2010/11 season. Similar results were shown in a simulation study [43], where larger RSV outbreaks were expected after non-pharmaceutical interventions were relaxed following the COVID-19 pandemic. A key implication of the association between the dynamics of susceptible populations and RSV transmission is that even a transient perturbation in susceptible populations could lead to a shifted pattern of disease transmission. This can be crucial for deploying and evaluating different vaccination strategies, and for assessing how vaccination coverage affects long-term disease patterns, contributing to informed decision-making in public health interventions.

## Materials and Methods

### Data sources

The data were obtained from microbiologically verified pediatric hospitalizations due to RSV and influenza virus at the Karolinska University Hospital. There was a consistent catchment area for the hospital between July 1998 to June 2018. We also obtained data on annual birth rates within the catchment area and weekly climate data, including temperature, precipitation, and air pressure.

### Demographic data

We used the *smooth*.*spline* function (with 10 degrees of freedom) implemented in R (version 4.3.2) to interpolate weekly birth rates. Within our transmission model, we divided the <1 year age class into 12-month age groups to more accurately capture aging among this age class. The remaining population was divided into 9 age classes: [1,2) years, [2,3) years, [3,4) years, [4,5) years, 5–9 years, 10–19 years, 20–39 years, 40–59 years and above 60 years old. We estimated the net rate of immigration/emigration for each age group (detailed in **S1 Text**) to produce a rate of population growth and age structure similar to that of northern Stockholm. Data on age-specific contact rates were obtained from [20] specifically, we used the POLYMOD contact matrix from the Netherlands which had a similar contact pattern as Sweden. Demographic data used in this study are publicly available on Github: https://github.com/keli5734/Sweden_study.

### Climatic data

The climatic variables used in this study were temperature (in Celsius) and relative humidity (as a percentage) from July 1998 to June 2018, and were available from on Github: https://github.com/keli5734/Sweden_study. Weekly averages were calculated from the daily data. To incorporate the climate data into the RSV transmission model, we normalized the data to between -1 and 1.

### The center of gravity and the intensity of RSV activity

The center of gravity of RSV activity for each season (*G*_*s*_) was measured as the mean epidemic week, with each week weighted by the weekly number of admissions (*Y* _*s,w*_), such that 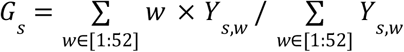, where *w* is an index for the week of each epidemic season, *s*. The intensity of RSV activity for each season was determined by the maximal values of hospitalizations.

### Wavelet analysis

We obtained the timing of RSV epidemics in each season based on phase decomposition obtained from wavelet analysis [44,45]. In the wavelet analysis, we used the 0.8–1.5 year periodicity band from the wavelet spectrum to extract weekly phases.

### Dynamic model description

Here, we used an age-stratified SIS (Susceptible-Infectious-Susceptible) model for RSV transmission dynamics. The model was proposed by Pitzer et al. [20] to study the environmental drivers of the spatiotemporal dynamics of RSV in the US. The model assumed individuals are born with protective maternal immunity, which wanes exponentially, leaving the infants susceptible to infection. We assumed a progressive build-up of immunity following up to four previous infections. Following infection with RSV, individuals develop partial immunity, reducing the rate of subsequent infection and relative infectiousness of the following infections. We also assumed subsequent infections have a shorter recovery time compared to the primary infection. The model was described by a system of ordinary differential equations (ODEs); see **S1 Text** for details.

### Parameter estimation

We first calibrated the transmission dynamic model for RSV to weekly RSV admissions from July 1998 to June 2008 (i.e., 11 seasons before the influenza pandemic). We estimated four parameters: a seasonal amplitude parameter (α), a seasonal offset parameter (ϕ), a baseline transmission rate parameter (β_0_), and a reporting fraction parameter (*f*). Note that the force of infection is given by *λ* =β_0_ (1 + α cos(2π*vt* − ϕ))*I*^*^, where *I*^*^ denotes all infection states. Two additional parameters were estimated when climate data were incorporated into the model: seasonal amplitude parameters for temperatures (α _*Temp*_) and relative humidity (α _*RelHum*_), such that *λ* =β_0_ (1 + α co*s*(2π*vt* − ϕ)+ α_*Temp*_ × *Temp* + α _*RelHum*_ × *RelHum*)*I*\* where *Temp* and *RelHum* are normalized data of temperatures and relative humidity. We estimated these parameters using a maximum likelihood approach, assuming the number of hospitalizations in each age class during each week was Poisson-distributed with a mean equal to the model-predicted number times the estimated reporting fraction. Other parameter values for the model were adopted from [20], and they are provided in **S1 Table**. We seeded the model with one RSV-infected individual in each age group except the <1 year group then used a burn-in period of 41 years to ensure the model reached a quasi-equilibrium steady state.

To estimate viral interference parameters, we applied a sampling-importance resampling method. We first used Latin Hypercube Sampling (LHS) to generate representative samples from a wide range of values for the parameter space ϕ =(ξ, η), where ξis a viral interference effect parameter, and η is a parameter for the duration of viral interference. We drew 100,000 samples from a uniform distribution *U*(−5, 0)for log _10_ (ξ), and from a uniform distribution *U*(1, 21)for η. Note that we drew samples using a log10-scale for the parameter ξbecause we had no prior knowledge of the magnitude of the parameter. Then, we generated forward simulations using the sampled parameter sets and fitted them to weekly RSV admissions from July 2008 to June 2018. We calculated the log-likelihoods of the model under each parameter set, assuming the number of hospitalizations in each age class during each week was Poisson-distributed with a mean equal to the model-predicted number times the estimated reporting fraction. We then normalized the log-likelihoods (as weights) of each parameter set and resampled 10,000 parameters from the joint distribution based on the weights.

## Data Availability

All data produced are available online at https://github.com/keli5734/Sweden_study

## Acknowledgments

This work was supported by a grant from the National Institutes of Health (R01AI137093). The content is solely the responsibility of the authors and does not necessarily represent the official views of the National Institutes of Health.

## Data and code availability

We used the R statistical software (v4.0.2) for all statistical analyses and visualization. Data and code used in this study are publicly available on Github: https://github.com/keli5734/Sweden_study

## Supplementary Materials

**Supplements Figure 1.**
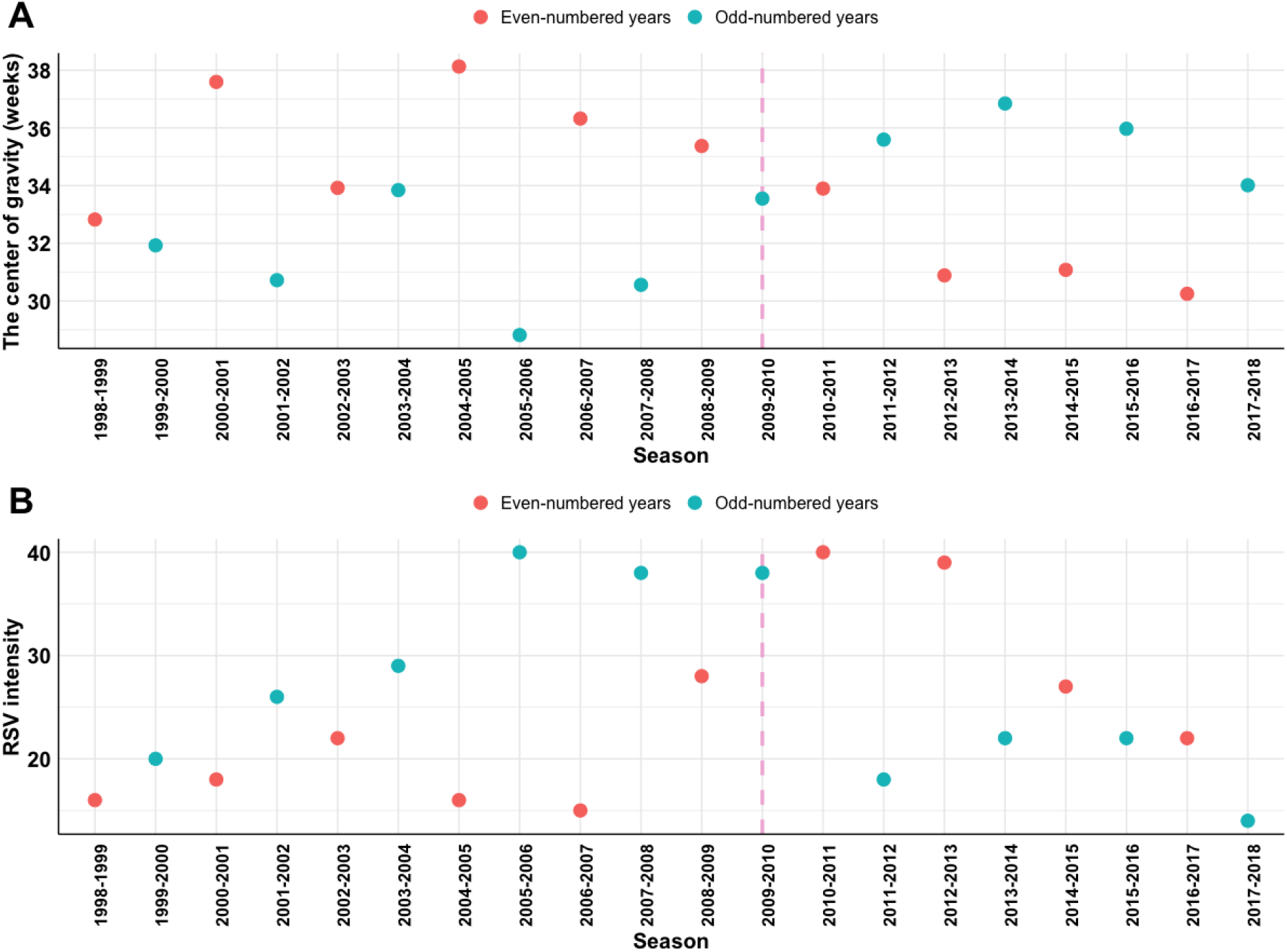
Measurements of RSV activity in northern Stockholm. **(A)** The mean timing (measured by the center of gravity, in week) of RSV epidemics. The vertical dashed line denotes the influenza pandemic 2009. **(B)** The intensity of RSV activity (measured by the peak value of RSV hospitalizations in each season). Colors indicate even-numbered (red) or odd-numbered (cyan) years.

**Supplements Figure 2.**
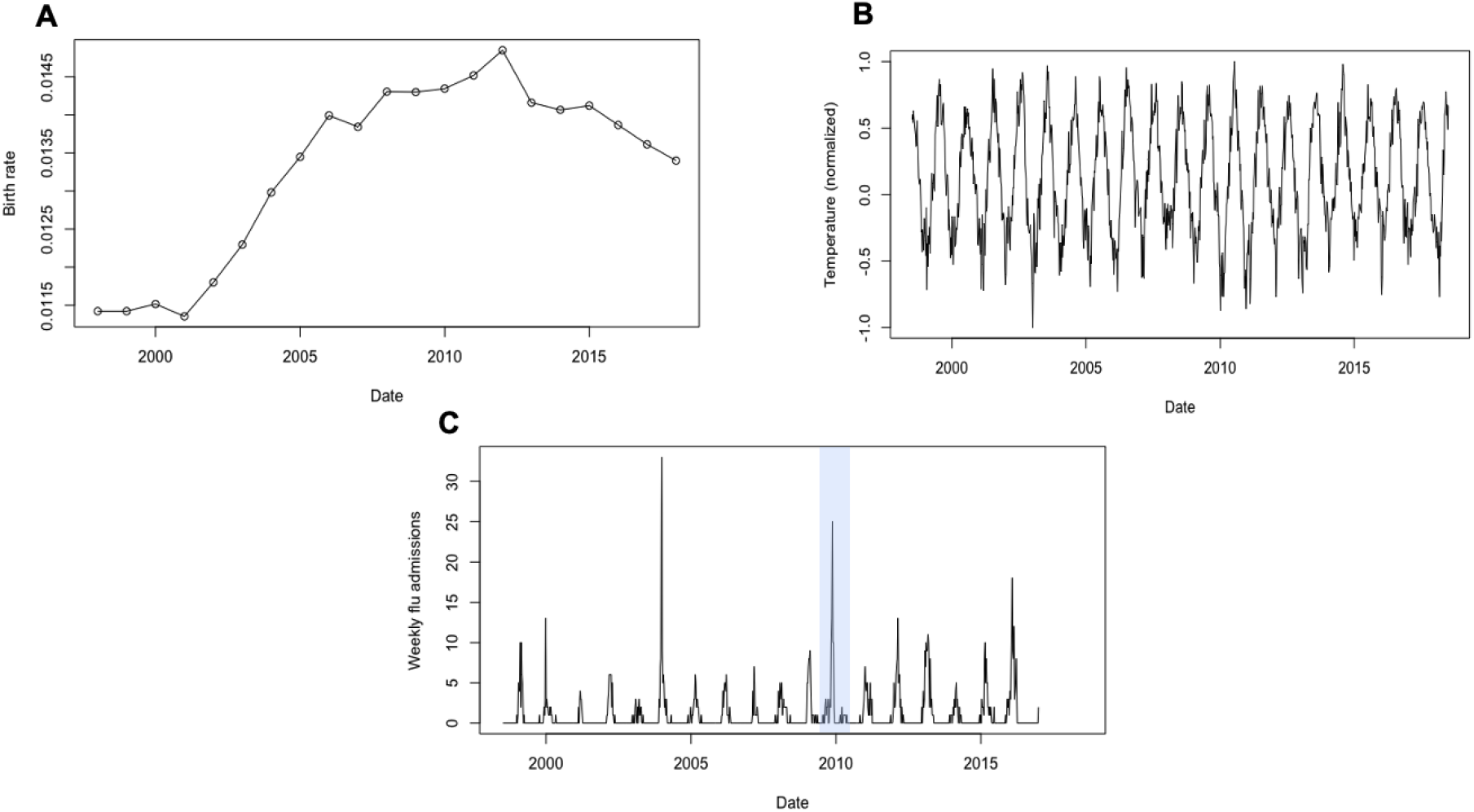
Time series data. **(A)** Annual Birth rates from 1998 to 2018 in northern Stockholm. **(B)** Weekly temperature (normalized) data. **(C)** Weekly admissions for influenza from 1998 to 2016. Note that we only used the data from pandemic seasons (highlighted in the shaded area) for model fitting.

**Supplements Figure 3.**
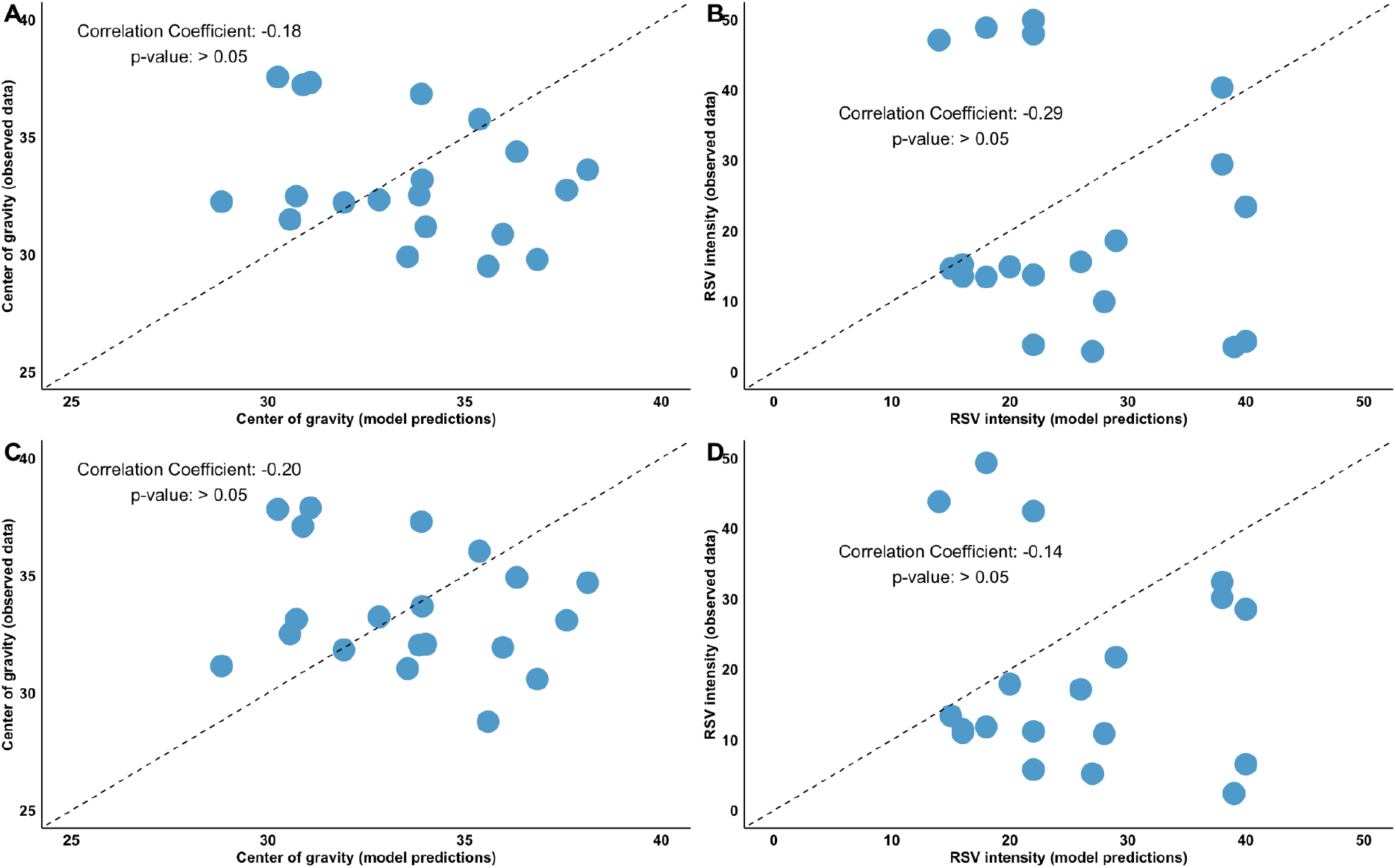
Time series data. Correlations between **(A)** the center of gravity (in weeks) and **(B)** RSV intensity measured by the peak value of RSV hospitalizations in each season of observed and model predicted RSV epidemics from the model only considering birth rates. Correlations between **(C)** the center of gravity (in weeks) and **(D)** RSV intensity measured by the peak value of RSV hospitalizations in each season of observed and model predicted RSV epidemics from the model considering birth rates and climate data.

**Supplements Figure 4.**
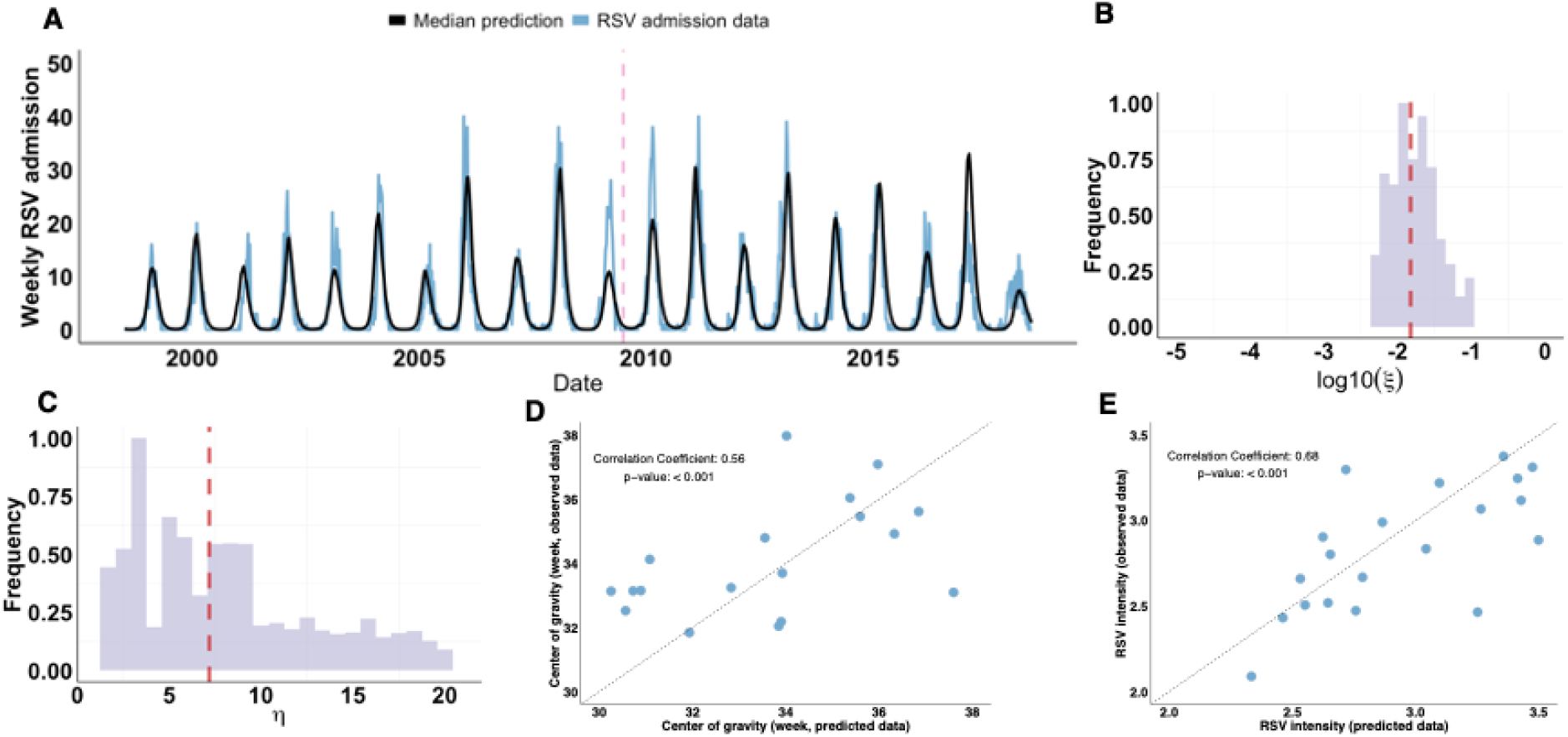
Model fit to age-stratified RSV admissions with viral interference effects and climatic factors. (**A**) The number of observed weekly RSV admissions are shown in blue; the median model prediction, given by the median estimates of viral interference parameters, is shown in black. The pandemic H1N1 influenza virus was introduced to the model only in the 2009/10 season, and the pink dashed line indicates July 2009. The marginal distribution for (**B**) the effect of viral interference parameter (ξ, in a log10-scale), and (**C**) the duration of viral interference parameter (η). The median estimates are indicated by red dashed lines (log_10_ (ξ)=−1. 82 and η =7. 18). Correlations between (**D**) the center of gravity (in weeks) and (**E**) the intensity of observed and predicted seasonal RSV epidemics from the best-fit model with the viral interference effect.

**Supplements Figure 5.**
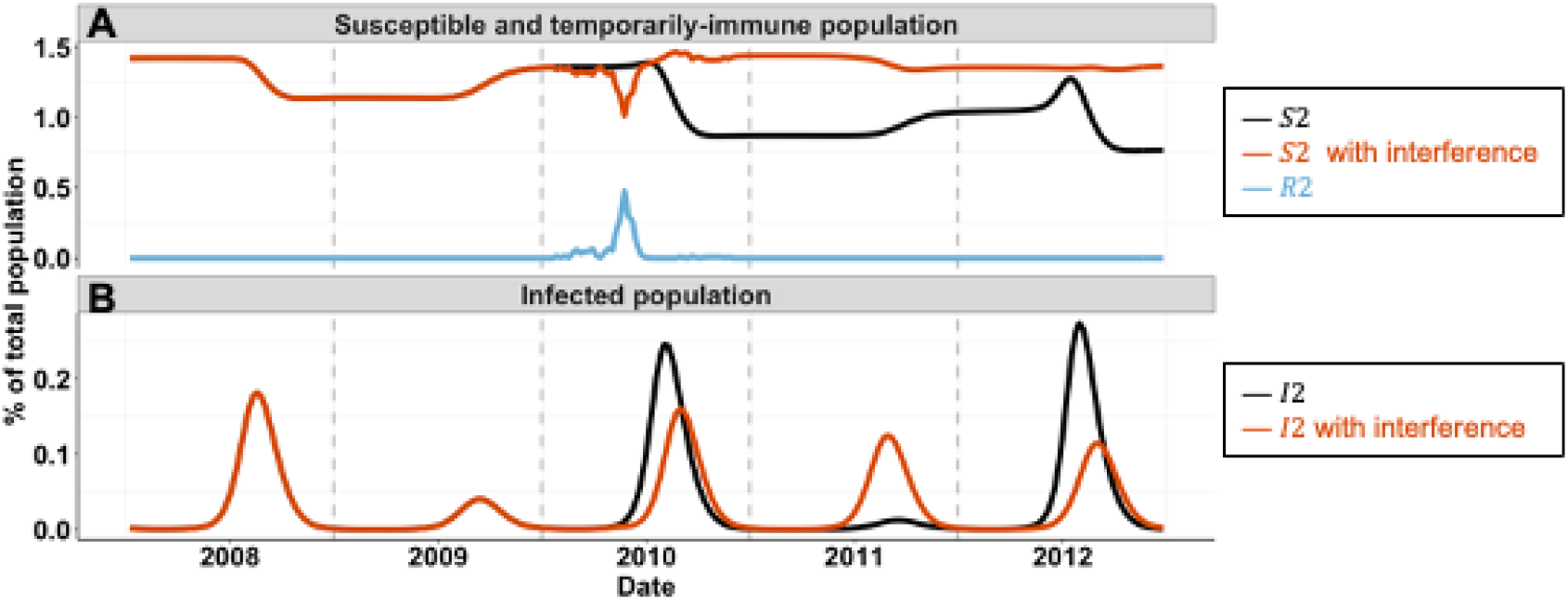
Model predictions of RSV epidemics following the influenza pandemic. (**A**) Predicted time series of recovered susceptible populations (*S*2) in the absence (black curve), or in the presence (red curve) of viral interference effects. The predicted time series of the temporarily immune population is shown in blue (*R*2). (**B**) Predicted time series of reinfected populations (*I*2) in the absence (black curve), or in the presence (red curve) of viral interference effects.

**Supplements Figure 6.**
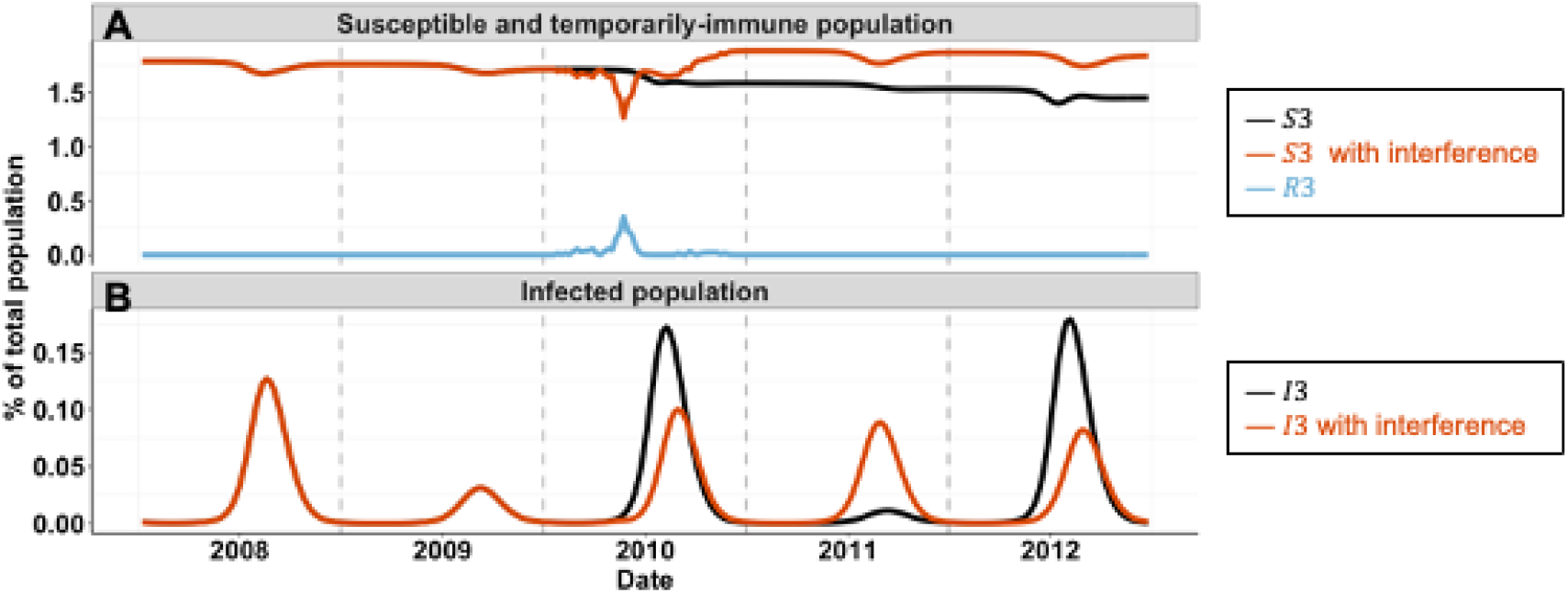
Model predictions of RSV epidemics following the influenza pandemic. (**A**) Predicted time series of recovered susceptible populations (*S*3) in the absence (black curve), or in the presence (red curve) of viral interference effects. The predicted time series of the temporarily immune population (*R*3) is shown in blue. (**B**) Predicted time series of reinfected populations (*I*3) in the absence (black curve), or in the presence (red curve) of viral interference effects.

**Supplements Figure 7.**
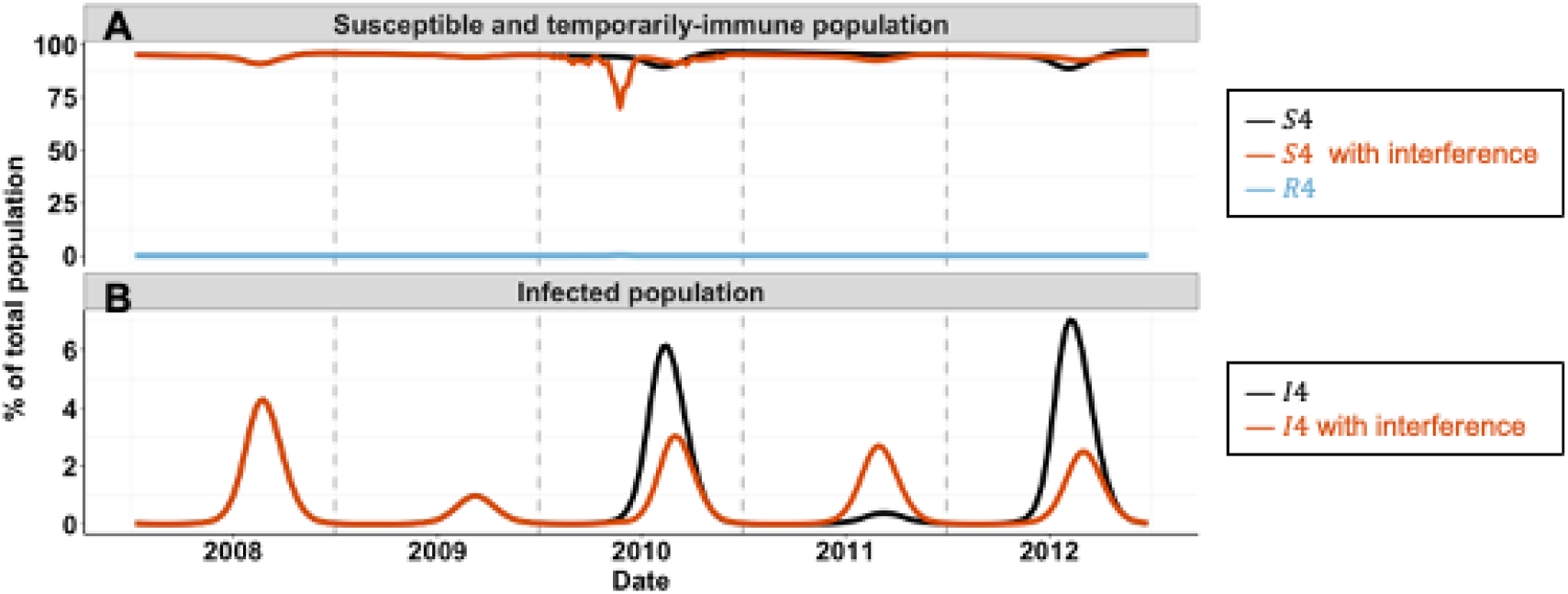
Model predictions of RSV epidemics following the influenza pandemic. (**A**) Predicted time series of recovered susceptible populations (*S*4) in the absence (black curve), or in the presence (red curve) of viral interference effects. The predicted time series of the temporarily immune population (*R*4) is shown in blue. (**B**) Predicted time series of reinfected populations (*I*4) in the absence (black curve), or in the presence (red curve) of viral interference effects.

**Supplements Table 1.** Transmission dynamic model parameters from Pitzer et al. [20].

| Parameter | Description | Parameter value |
| --- | --- | --- |
| $1/\omega$ | Duration of maternal immunity | 16 weeks |
| $1/\gamma_1, 1/\gamma_2, 1/\gamma_3$ | Duration of infectiousness | 10 days, 7 days, 5 days |
| $\sigma_1, \sigma_2, \sigma_3$ | Relative risk of infection | 0.76, 0.6, 0.4 |
| $\rho_1, \rho_2$ | Relative infectiousness | 0.75, 0.51 |
| $d_{p,0}, d_{p,0.5}, d_{p,1}, d_{p,2}, d_{s,a}$ | Proportion of infections leading to LRT infection (First infection < 6 months, 6-11 months old, 1-2 years old, >2 years old, second infection) | 0.5, 0.3, 0.2, 0.1, $0.75*d_{p,a}$ |

**Supplements Table 2.** Estimated model parameters without viral interference effects and climatic data.

| Parameter | Description | Parameter value |
| --- | --- | --- |
| $\beta_0$ | Baseline transmission rate | 8.52 |
| $\alpha$ | Seasonal amplitude | 0.27 |
| $\phi$ | Seasonal offset | 3.24 |
| $f$ | Reporting fraction | 0.45 |

**Supplements Table 3.** Estimated model parameters including climatic data, without viral interference effects.

| Parameter | Description | Parameter value |
| --- | --- | --- |
| $\beta_0$ | Baseline transmission rate | 9.14 |
| $\alpha$ | Seasonal amplitude | 0.26 |
| $\alpha_{Temp}$ | Seasonal amplitude for temperatures | 0.09 |
| $\alpha_{RelHum}$ | Seasonal amplitude for relative humidity | 0.05 |
| $\phi$ | Seasonal offset | 3.30 |
| $f$ | Reporting fraction | 0.28 |

## Supplements Text 1

### Model equations

The set of differential equations describing the transmission dynamics of RSV is given as follows:

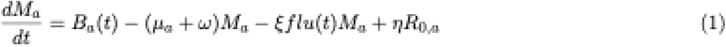

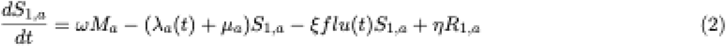

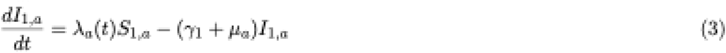

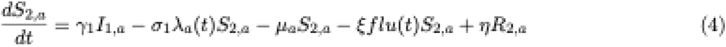

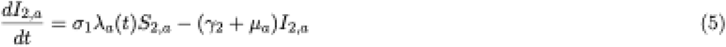

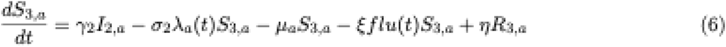

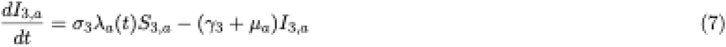

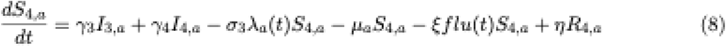

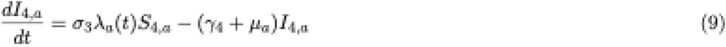

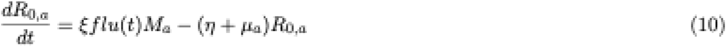

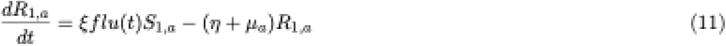

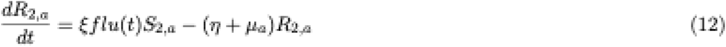

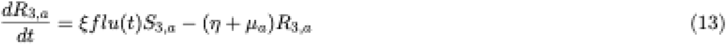

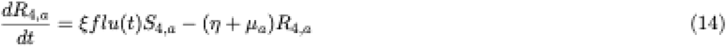

The force of infection of RSV for the age group a is *λ*_*a*_ (*t*), and they are given by

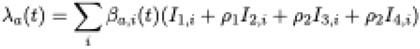

where 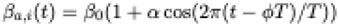.

We assume that only first and second infections can result in sever lower respiratory disease (*D*), and cases reported in the hospitalization and laboratory data (*H*_*a,w*_) are proportional to *D*_*a,w*_ The probability (*d*_*n,a*_) is dependent upon both age and number of previous infections. See Table Si for parameter values. Note that *D* and *H* represent disease states and not infection states, and therefore are not included in the differential equations above.

### Estimating net rates of iinmigration/emigration

We used *smooth*.*spitne* function (with 10 degrees of freedom) implemented in R (version 4.3.2) to interpolate the birth rates to calculate weekly birth rates. We divided the <1 year age class into 12—month age groups to more accurately capture aging among this age class. The remaining population was divided into 9 age classes: |1,2) years. [2,3) years, |3,4) years, [4,5) years, 5 9 years, 10 19 years, 20 39 years, 40 59 years and above 60 years old. We estimated the net rate (*μ* _*a*_) of immigration/emigration for each age group (*N*_*a*_) to produce a rate of population growth and age structure similar to that of north Stockholm, according to the equations:

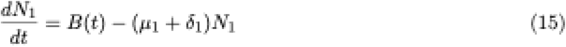

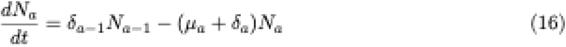

where *δ*_*a*_ is the aging rate (per week) in each age group (*a*) and equals to 1/(width of the age class).

**Figure S1.**
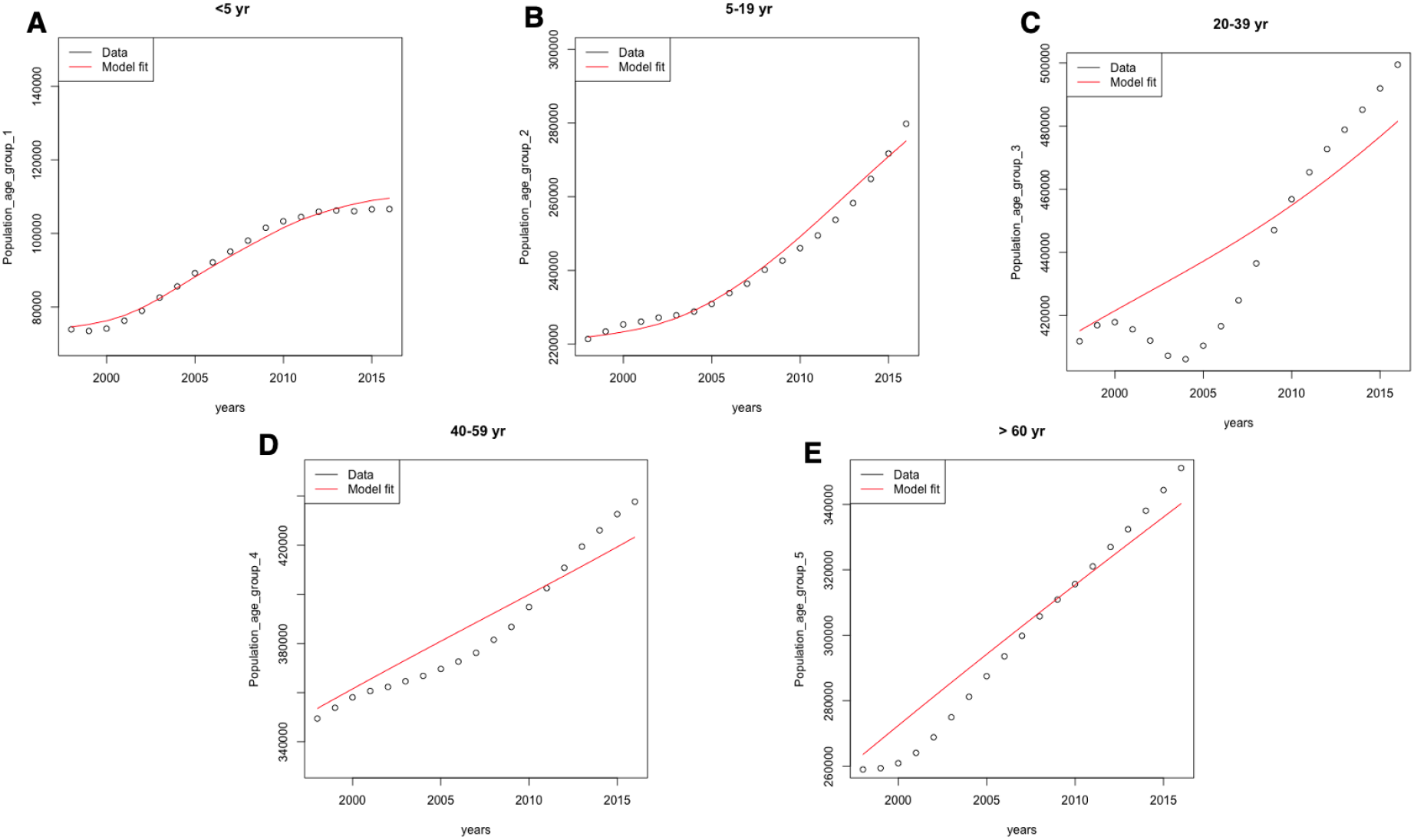
Model fit to demographic data.

